# The Impact of Marital Status and Sex on Heart Failure Readmissions: A Case Study from Atrium Health Floyd Medical Center

**DOI:** 10.64898/2025.12.23.25342948

**Authors:** Shriya Garg, Perisa Ashar, Brandi Skeen, Keely Harris, Stephanie Durall, Khushi Kohli, Rahul Garg

## Abstract

**Intro:** Heart failure is one of the major causes of death in the United States, characterized by high readmission rates annually. Our study sought to analyze de-identified patient data from the Atrium Health Floyd Medical Center to identify differences in readmission rates for patients based on sex and marital status and propose novel solutions to our findings.

**Methods:** In this retrospective cohort analysis, 1,122 HF patient discharges (obtained by reviewing heart failure specific diagnosis-related group and principal diagnosis codes) between February 1, 2023 – March 30, 2024 were analyzed to describe demographic trends of readmission and total length of stay. Such analyses included a chi-square test of independence and Kruskal Wallis Test alongside the appropriate post-hoc analyses. Univariable logistic regression analyses were used to understand the odds of readmission amongst the different groups of marital status and sex as well as between different rural/urban statuses. Regressions were visualized with forest plots.

**Results:** The heart failure readmission rate was 13.25% (147/1,109). Prior to post-hoc analyses, there was a significant difference in the observed count and expected count of married female readmission (9 vs. 19.161, p=0.020), and two proportion tests revealed a significantly higher readmission rate for single females compared to married females [14.36% (56/390) vs. 6.21% (9/145); p=0.010]. Prior to correction, married females had a statistically longer total length of stay compared to single females (p=0.038). Lastly, univariable logistic regressions revealed that married males, single females, and single males all had significantly higher odds of being readmitted compared to married females (married male: OR=2.248, p=0.045; single female: OR=2.532, p=0.013; single male: OR=2.606, p=0.010). No significant relationship between marital status and sex with length of stay or between geographic classification (metropolitan, micropolitan, rural, small town) with readmission was found.

**Conclusions:** Married females had the lowest readmission risk, while all others experienced significantly higher odds of readmission, pointing to the potentially protective role of partner-based social support post-discharge. Length of stay was not significantly related to marital or sex groups after correction suggesting that inpatient care delivery may be less sensitive to sociodemographic factors than post-discharge recovery. These findings underscore the need for better transitional care strategies for patients who lack strong support networks at home and stronger research backing to further characterize heart failure readmissions.

## INTRODUCTION

Heart disease, including diseases of the heart and blood vessels such as ischemic heart disease, stroke, congestive heart failure, hypertension and atherosclerosis, is the leading cause of death for Americans, with over 919,032 deaths due to cardiovascular disease in 2023, with 425,147 (45%) of these deaths due to heart failure (HF).^1,2^ In Georgia, cardiovascular disease results in over 28,000 deaths a year, with 165,103 years of potential life lost per year in Georgia.^3^ Magnifying the burden of heart-related conditions on patients, nearly 1 in 4 HF patients are readmitted within 30 days of discharge and approximately half within 6 months,^4^ a quarter of which were altogether preventable. In total, some projections indicate that over $858 billion dollars may be spent on managing HF by 2050.^5^ In summary, HF is becoming an increasingly common principal discharge diagnosis while also demonstrating high numbers of readmission.^6^

Heart failure readmission (HFR) is extremely costly both physically and mentally for patients. Recurrent HFRs are associated with increased risk of cardiovascular mortality (i.e. death resulting from diseases affecting the heart and/or circulatory system) and a lower health-related quality of life.^7–10^ Furthermore, increased hospital stays can dramatically increase the financial cost and burden on both the patient and their family, known as financial toxicity. Such financial distress due to medical costs, affecting over 137 million (56%) Americans nationwide,^11^ can lead to delayed or forgone care and lack of adherence to medication, both of which could worsen health outcomes.^12^ Furthermore, financial toxicity is associated with increased emotional burden (for the patient and caregiver), worries about affording other essentials such as food or accommodations, health-worsening stress, and more.^13^

Readmission into the hospital is not only detrimental to patients, but they are harmful to hospital profits as well. This is largely due to The Hospital Readmissions Reduction Program (HRRP), launched by the Centers for Medicare and Medicaid Services, which is a Medicare value-based purchasing program designed to incentivize the reduction of avoidable readmissions. The program links hospital Medicare reimbursements to the number of readmissions, with higher readmissions reducing payment by up to 3%.^14^ In Rome, GA, where this study was performed, the hospitals in the area were also subject to HRRP. Floyd Medical Center, now Atrium Health Floyd, lost 0.32% of Medicare reimbursement dollars.^15^ On average, each patient readmission costs $15,200, ranging from $10,900 for self-pay/no charge stays to $16,400 for patients with private insurance.^16^

In this study, we sought to characterize the demographic characteristics (such as marital status and sex) to identify potential associations with readmission and total length of stay for HF patients.

## METHODS

### Data Set

We performed a retrospective cohort analysis of 1,122 HF patient discharged between February 1, 2023 – March 30, 2024, to describe differences in heart failure readmission rates based on sex and marital status and identify novel solutions to reduce readmission burden on patients and healthcare facilities. Deidentified patient data was obtained through retrospective audits of data collected on patients with the principal diagnosis codes related to HF at Atrium Health Floyd Medical Center.^17^ The study required and obtained institutional board review as it constitutes human subject research.

In this study, we categorized all patients diagnosed with HF by utilizing diagnosis-related group codes (DRG) including 287, 291, 292, 293, 280, 264, 286, 246, 226, 247, 281, 291, 255, 283, 252, 258, 242, 282, 273, 250, 004, 321, 194, 276, 192 (**Supplementary Table 1)**. Principal diagnoses codes included in this study include: I11.0, I50.33, I13.0, I50.43, I13.2, I50.23, I50.21, I50.31, and I50.41 (**Supplementary Table 2).** Diagnosis codes 004 and 194 were included because those patients received a principal diagnosis code related to heart failure.

The primary outcome was the readmission. Readmission, in this study, is defined as admissions to an acute care hospital within 30 days of discharge for any condition or diagnosis.^18^ We investigated differences in readmission and length of stay amongst self-identified sex (female or male) and marital status. We examined heart failure readmission differences across combinations of marital status and sex. Patients with unknown sex and marital status were excluded from analyses. Race and ethnicity variables were collected from the medical record but were not analyzed because of small subgroup sample sizes and limited diversity in the study population.

### Data Processing and Statistical Methods

To perform these analyses, we created two groups of patients, those in the single grouping (i.e. single, divorced, widowed, or legally separated) and those in the married grouping. We conducted a Chi-square test of independence and Post hoc (using an Benjamini-Hochberg (BH) procedure) to identify any associations between marital status and sex with readmission. From there, we identified an interesting association between married female and single female readmission and conducted a two-proportions test. To identify associations between marital status and sex with length of stay, we conducted a Kruskal-Wallis Test across the groups. Subsequent post-hoc Dunn tests (using Benjamini-Hochberg correction) were conducted to evaluate any statistically significant differences for pairwise comparisons.

Furthermore, we conducted univariable logistic regression analyses. Adjusted odds ratios and 95% confidence intervals of heart failure readmission were measured for each of the groups (single female, married male, single male) compared to the reference group (married female). From this, P-values were calculated using the alpha level of 0.05. Similar univariable logistic regressions were conducted to analyze the relationship between metropolitan, micropolitan, small town and rural area geographic classifications, as defined by primary Rural-Urban Commuting Area Codes (RUCAs) and readmission. RUCAs help to differentiate between rural and urban areas throughout the U.S., and primary RUCAs help to establish urban cores and the census tracts that are the most economically integrated with those cores through commuting.^19^ To visualize our results, we utilized forest plots and mapping in RStudio (Version 2025.09.2+418). Multivariable adjustment was not performed because (1) sex was incorporated directly into the exposure definition and therefore not suitable for adjustment, and (2) key potential confounders such as cardiac function, comorbidity burden, and outpatient access were insufficiently captured to support reliable multivariable modeling.

## RESULTS

From this study, we found that the total HFR rate for this data set, limited to between February 1, 2023, and March 30, 2024, was 13.19% (148/1,122). The mean age for patient discharges in this data set was 65.97, with married females averaging 67.9 years of age, married male averaging 70.16 years of age, single females averaging 67.6 years of age, and single males averaging 62.0 years of age. From our patient data set, 789 (70.3%) were from metropolitan areas, 238 (21.2%) patients were from Micropolitan areas, 56 (4.99%) patients were from small towns, and 39 (3.48%) were from rural areas (**Table 1**).

**Table 1:**
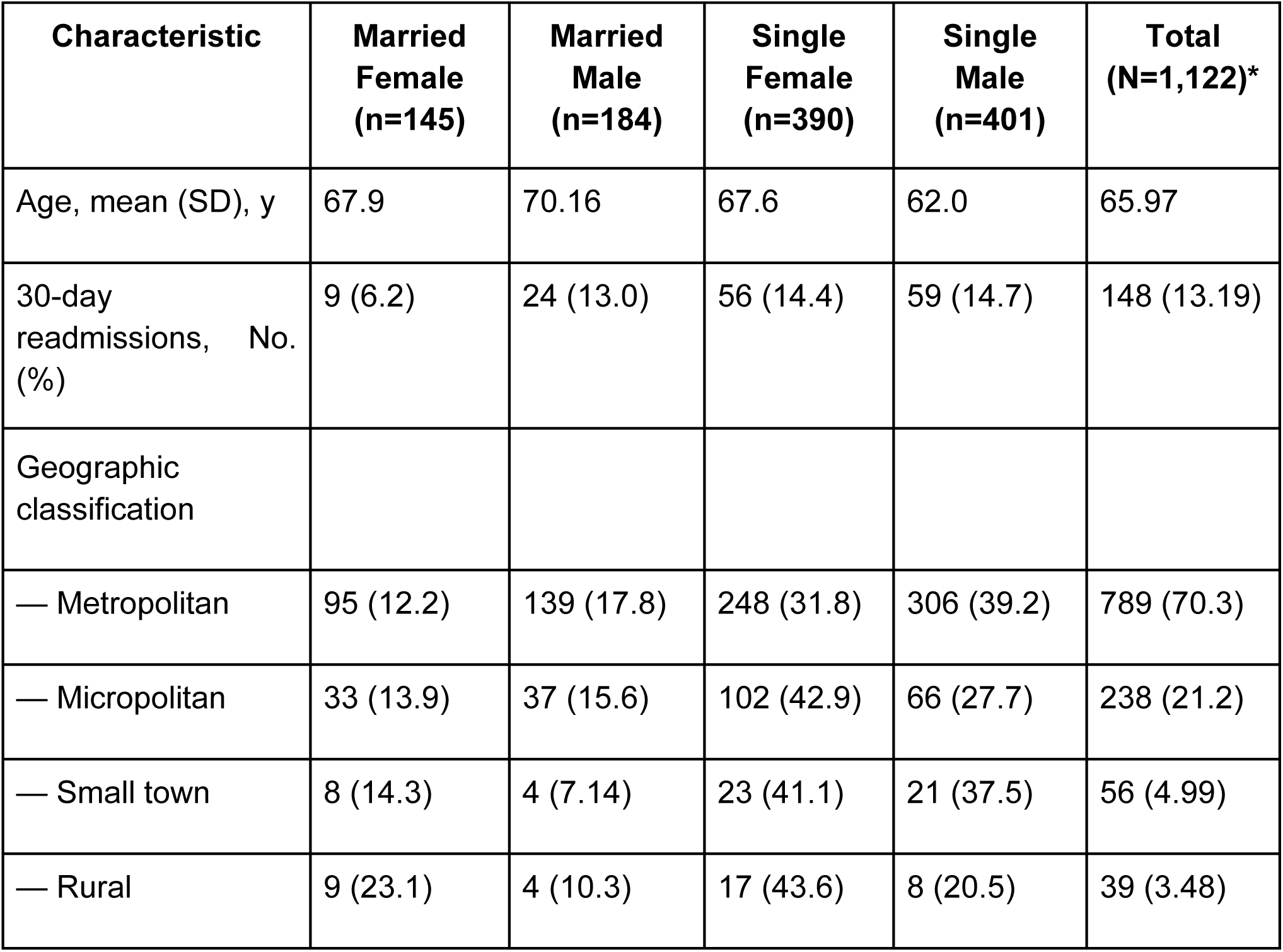
Baseline Characteristics of Patients Hospitalized for Heart Failure, by Sex and Marital Status.

Our Chi Square Test of Independence (global p-value 0.059) confirmed that the observed count of married female readmission was statistically different (prior to corrections) from the expected count of married females’ readmission (9 vs. 19.161, P=0.020) (**Supplementary Table 3**). However, Dunn’s post hoc tests with Benjamini-Hochberg corrections led to a p-value of 0.162 (**Supplementary Table 4)**. Because the comparison between married and single women represented a predefined, clinically meaningful contrast regarding the role of partner-based support in recovery, we conducted a targeted two-proportion test to directly evaluate differences in event rates. A two-proportion test was selected because it more directly quantifies differences in readmission percentages between two independent groups without relying solely on omnibus chi-square assumptions. This analysis confirmed that single women experienced significantly higher 30-day readmission compared with married women [14.36% (56/390) vs 6.21% (9/145); P=0.010].

Our Kruskal-Wallis Test Across Groups revealed that there were statistically significant associations between marital status and sex (P=0.041). Per Dunn’s post-hoc tests, married females had a statistically longer total length of stay compared to single females (P=0.038) prior to correction (P=0.084). Furthermore, married females had a statistically smaller total length of stay compared to married males (P=0.035) prior to correction (P=0.084) (**Supplementary Table 5).**

Our mapping revealed that the highest counts of readmission for patients with the specified DRG codes were those counties that were closest to the Atrium Health Floyd. The counties that were further on the perimeter from the hospital reported 0 readmissions, likely with patients being readmitted to other areas (**Figure 1).**

**Figure 1:**
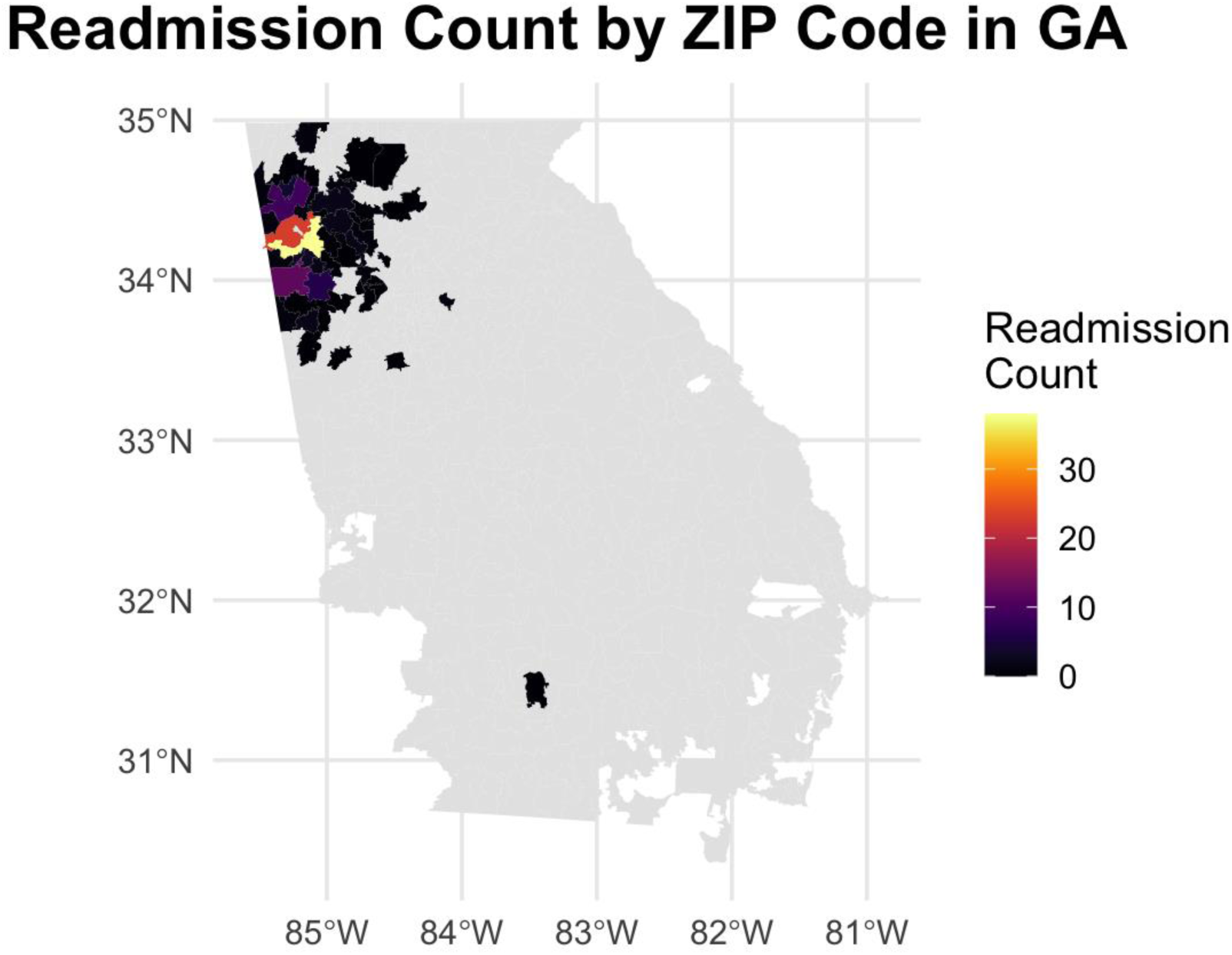
Depiction of Total Readmission Count by Zip Code in Georgia

Univariable logistic regressions revealed that married males, single females, and single males all had significantly higher odds of being readmitted compared to married females (married male: OR=2.248, P=0.045; single female: OR=2.532, P=0.013; single male: OR=2.606, P=0.010) (**Supplementary Table 6**) (**Figure 2).** Univariable linear regressions revealed no significant relationship between marital status and sex with length of stay (**Table 2) (Figure 3)**. Additionally, univariable linear regressions revealed no significant relationship between geographic classification (metropolitan, micropolitan, rural, small town) with readmission.

**Figure 2:**
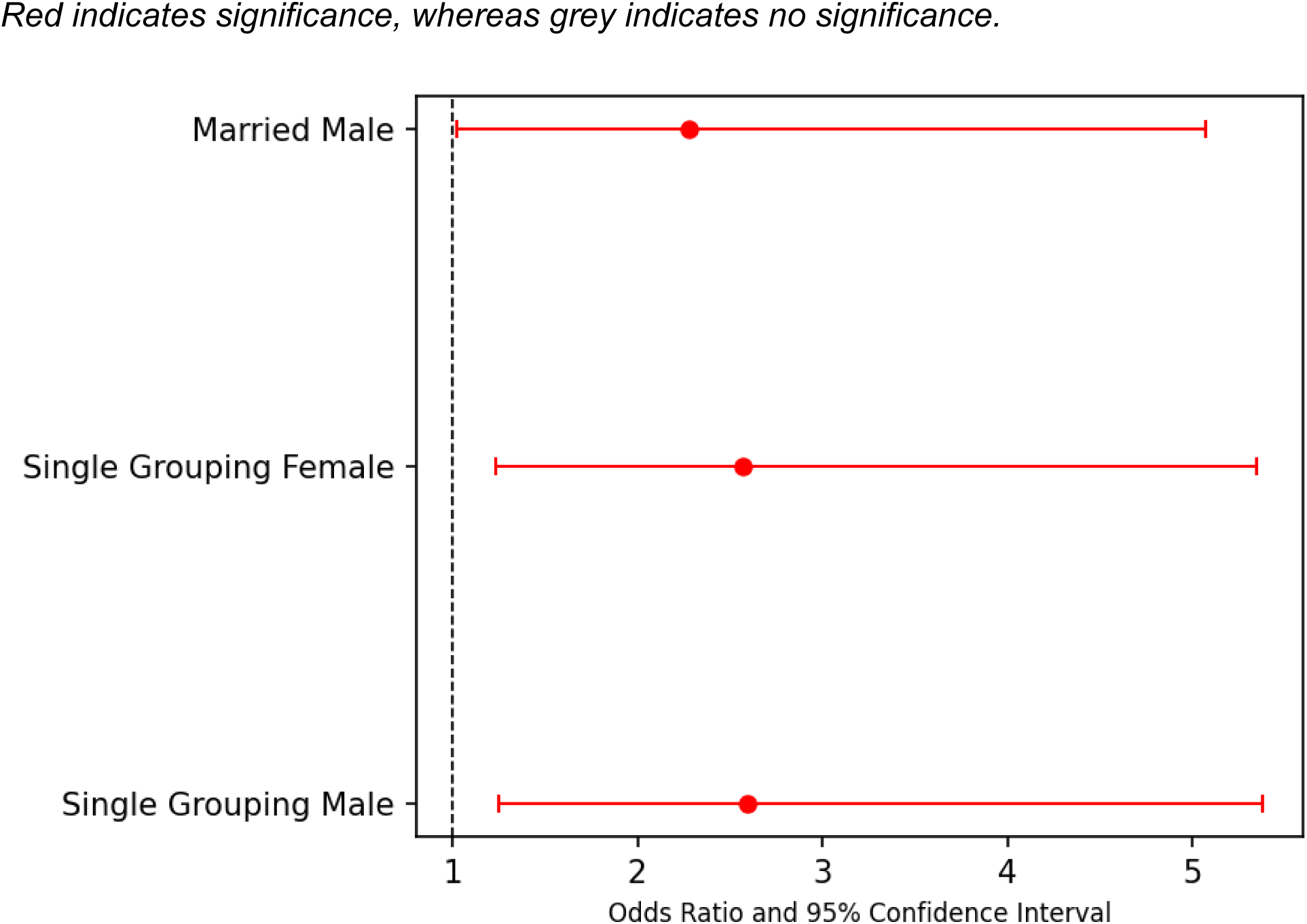
Forest Plot visualizing Odds Ratio and 95% Confidence Interval for Readmission by Marital Status and Sex (Reference: Married Females)

**Figure 3:**
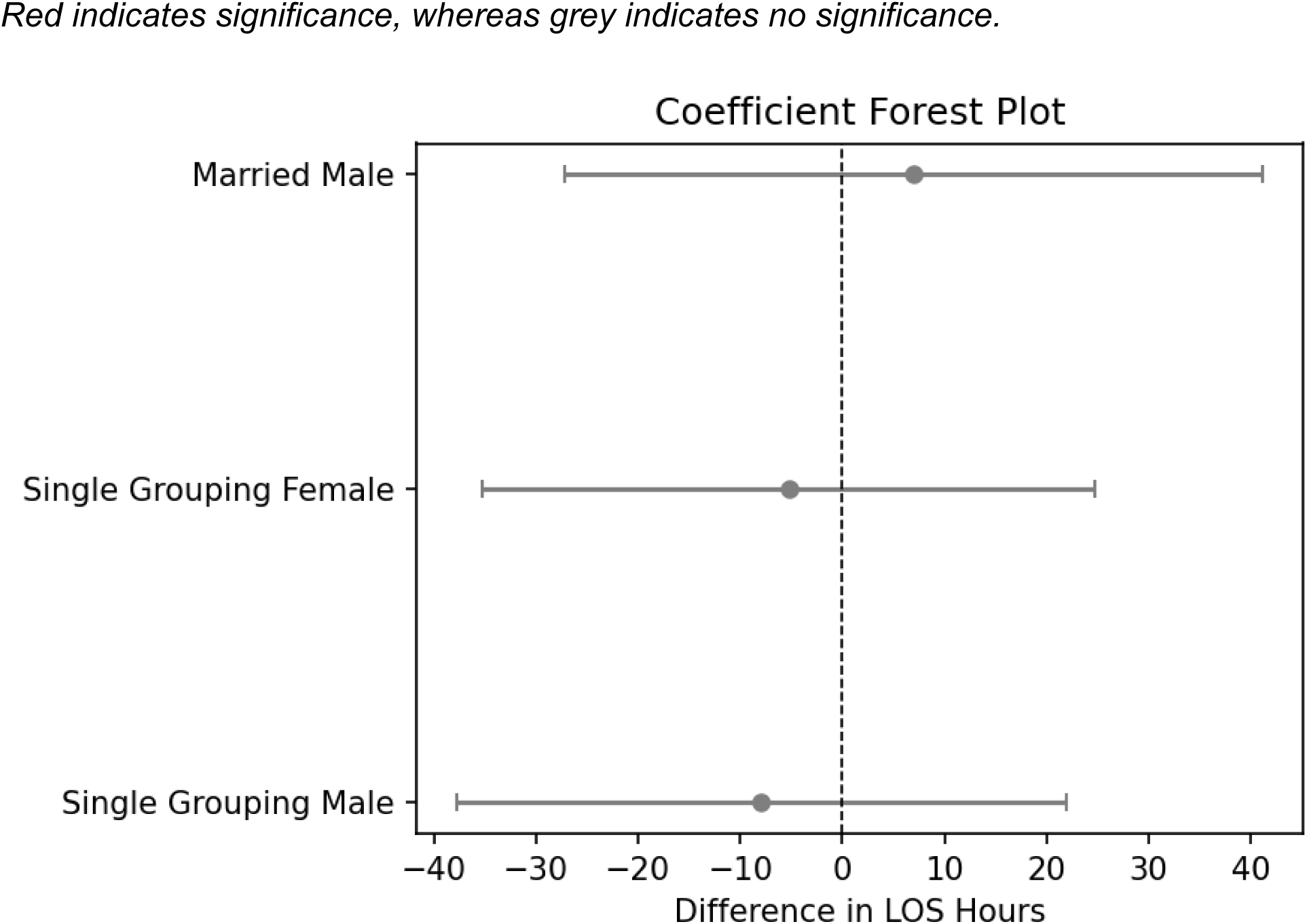
Forest Plot visualizing Regression Coefficients and 95% Confidence Interval for Total Length of Stay by Marital Status and Sex (Reference: Married Females)

**Table 2:** Heart Failure Readmission Risk and Length-of-Stay Differences by Marital Status and Sex *(Reference group = Married Female)*

| **Group** | **30-Day**<br>**Readmission %**<br>**(n/N)** | **Odds Ratio**<br>**(95% CI)\*** | **p-value\*** | **LOS Difference**<br>**vs Married**<br>**Female† (95% CI)** | **p-value†** |
| --- | --- | --- | --- | --- | --- |
| **Married**<br>**Female** | 6.2% (9/145) | Reference | — | Reference | — |
| **Married**<br>**Male** | 13.0% (24/184) | OR = **2.25**<br>(1.02–4.96) | **0.045** | **+7.00** hours | 0.688 |
| **Single**<br>**Female** | 14.4% (56/390) | OR = **2.53**<br>(1.22–5.17) | **0.013** | **-5.26** hours | 0.731 |
| **Single**<br>**Male** | 14.7% (59/401) | OR = **2.61**<br>(1.26–5.41) | **0.010** | **-7.93** hours | 0.603 |

\*From univariable logistic regression
†From univariable linear regression for length of stay

## DISCUSSION

HF remains a leading cause of hospitalization, readmission, and death in the United States, increasing average healthcare costs and potential morbidity and mortality for patients, while reducing patient and family quality of life.^20^ Reducing 30-day readmissions is essential to improving the patient care delivery, and programs like HRRP help to align the incentives of hospitals to do so. However, through the identification of demographic groups at higher risk for readmission and advocating for better transitional care, community intervention, and heart failure education, hospitals can help to alleviate the burden of HF readmission on patients and hospitals alike.

In this study, we sought to understand demographic characteristics that might predict differences in heart failure-related readmission and/or total length of in-patient stay while also conducting analyses on geographic location as well. We did so to highlight potential subpopulations that might benefit from more supportive and targeting programming to reduce the burden of heart failure and potential readmission on patients, families, and hospitals. We found that married females (i.e. patients that self-identified as female and were married) were less likely to be readmitted within 30 days of discharge compared to single females (i.e. patients who were single, divorced, widowed, or legally separated). Moreover, married males, single females, and single males all had significantly higher odds of readmission compared to married females.

These findings are consistent with existing literature, which found that younger patients with acute myocardial infarction between the ages of 18-55 were more likely to be readmitted within 1 year of discharge if they were unpartnered.^21^ Another such study found that being married and living with other family members independently predicted lower heart-failure related mortality and fewer readmissions for African American patients.^22^ Finally, one study which derived its results from the ACS Israeli Survey, found that marriage is associated with better short- and long-term outcomes and advocated for greater prevention measures for unmarried patients.^23^ This suggests that there are potential health benefits associated with having a partner, including a reduced likelihood of being readmitted into the hospital for cardiovascular-related concerns.^24^

Health benefits associated with having a partner can include greater operational social support (medication management and appointment upkeep),^25^ better treatment, symptom monitoring and quality of life,^26^ symptom monitoring, more access to transportation for appointments, protection from psychological distress or depressive symptoms,^27^ and more.^28^ Alternatively, having a partner could help to build economic capital, supporting healthier behaviors and lifestyles. The difference in readmission between patients of differential marital status reveals the potential for marriage protection and selection, or the idea that healthy patients are more likely to have a partner.^29,30^ This theory can be reflected in our results with the higher readmission for single males and females, despite the longer total length of stay of married females compared to single females.

However, our findings were unique due to our focus on both marital status and sex associations with readmission which are still unclear in the literature. Studies on difference in sex, in combination with the impact of partner status on readmissions, have led to mixed results, with studies varying vastly on whether females or males experience higher readmission rates and few focusing on combined marital and sex status.^31^ Those studies that focus on sex have largely found no statistical relationship between married females and males, in terms of outcomes and readmissions.^32,33^ These findings underscore the need to further evaluate disparities in heart failure readmission.

We additionally found no significant relationship between geographic classification (metropolitan, micropolitan, rural, small town) with readmission. Such findings were surprising to the insurmountable amount of evidence that patients from rural, underserved areas typically have higher 30-day readmission rates.^34,35^ However, from our study, we also observed geographic clustering of heart failure readmissions near Atrium Health Floyd, with lower readmission counts from surrounding areas on the periphery. Such patterns may reveal geography-centered care seeking, where patients seek care at hospitals that are closer in distance to them, rather than giving us insight into potential differences in local healthcare access and utilization. This highlights the need for future research and data that links patient data from many hospitals (not just Atrium Health Floyd) allowing for the heart failure readmissions for the entire local population.

Our study highlights the key question “How do we provide the social support and resources that female married patients have to reduce readmission rates among other heart failure patients?” We have underscored the need for more target programming to hone in on the protective aspects of partnership and marriage. One type of targeted programming includes enhanced transitional care. Existing literature strongly supports the success for early outpatient follow up after discharge, as patients who are discharged from hospitals and have early follow-up rates have lower risk of 30-day readmission likely due to greater clinical monitoring and treatment compliance.^36,37^ Another meta-analysis found that early post-discharge clinical follow up led to a 22% reduction in readmission risk.^38,39^

Alternatively, post-discharge telephone calls are another low-barrier transitional care option. One study found that structured telephone visits after hospitalization were more likely to increase 7-day follow-up and reduce in-person visits,^40^ while another randomized controlled trial found that patients who received weekly phone calls for 30-days post-discharge had 44% lower readmission rate.^41^ Virtual or over-the-phone care coordination helps to build stronger patient-clinician relationships while also being more cost- and time-effective.^42^ Other potential interventions, including comprehensive disease-management programs (tailored to those who are single), patient education about heart failure risk factors, and social needs screening may help to reduce the burden of heart failure nationwide. Lastly, more conversations with clinicians about at-home social support could help physicians and nurses identify the patients in need of more specific heart failure-related care.

Our study has many limitations. First, due to the small sample size and lack of diverse race/ethnicity data (due to the composition of the area in which Atrium Health Floyd is located likely), we were unable to conduct a robust race and ethnicity analysis for heart failure readmission rates – despite the large amount of evidence pointing to differences in readmission and mortality for patients of minority status. Secondly, due to our focus on marital status, we grouped all unmarried patients into the single category. In the future, we believe that a separate study might reveal differences in readmission and total length of stay for patients who were divorced, widowed, never married, and more. Furthermore, all the data used in this study was self- reported and, thus, may be subject to reporting bias. For example, patients may underreport risk factors such as drinking, smoking, poor diet, exercise and more – all of which are contributors to heart disease and failure.^43^ Because limited clinical covariates were available in this retrospective dataset, univariable models were used to estimate unadjusted associations and generate hypotheses for future study. Future multivariable analyses incorporating measures of cardiac severity, comorbidity burden, socioeconomic context, and outpatient access are needed to determine whether the protective effect of partner-based support persists after accounting for clinical risk factors.

## CONCLUSIONS

In this retrospective single-center study of more than 1,100 patients hospitalized for heart failure, marital status and sex were meaningfully associated with 30-day readmission, whereas rural and urban geographic classification demonstrated no significant effect on readmission or length of stay. Married females had the lowest readmission risk, while single males, married males, and single females all experienced significantly higher odds of readmission. These results highlight the potentially protective role of partner-based social support during the post-discharge period. Length of stay did not significantly differ across marital or sex groups after correction, which suggests that inpatient care delivery may be less sensitive to sociodemographic factors than post-discharge recovery.

These findings support the importance of targeted transitional care strategies for patients who lack strong support networks at home. Future studies should incorporate broader clinical determinants of readmission, such as cardiac function, comorbidities, and access to timely outpatient follow-up, and should leverage multi-center datasets to capture readmissions occurring outside the index hospital. Tailoring heart failure interventions to socially vulnerable patient groups may reduce avoidable readmissions and improve both patient outcomes and health system performance under value-based care programs.

## Data Availability

Deidentified individual participant data that underlie the results reported in this article will not be made available. The data were obtained from a single health system and are subject to institutional review board restrictions and data use agreements that do not permit public data sharing.

## Supplementary Materials

**Supplementary Table 1:** A List of Unique Diagnosis-Related Group (DRG) Codes Used.

| DRG Code | Definition |
| --- | --- |
| 287 | CIRCULATORY DISORDERS EXCEPT AMI, WITH CARDIAC CATETERIZATION WITHOUT MCC |
| 291 | HEART FAILURE AND SHOCK WITH MCC |
| 292 | HEART FAILURE AND SHOCK WITH CC |
| 293 | HEART FAILURE AND SHOCK WITHOUT CC/MCC |
| 280 | ACUTE MYOCARDIAL INFARCTION, DISCHARGED ALIVE WITH MCC |
| 264 | OTHER CIRCULATORY SYSTEM O.R. PROCEDURES |
| 286 | CIRCULATORY DISORDERS EXCEPT AMI, WITH CARDIAC CATHETERIZATION WITH MCC |
| 246 | PERCUTANEOUS CARDIOVASCULAR PROCEDURES WITH DRUG-ELUTING STENT WITH MCC OR 4+ ARTERIES OR STENTS |
| 226 | CARDIAC DEFIBRILLATOR IMPLANT WITHOUT CARDIAC CATHETERIZATION WITH MCC |
| 247 | PERCUTANEOUS CARDIOVASCULAR PROCEDURES WITH DRUG-ELUTING STENT WITHOUT MCC |
| 281 | ACUTE MYOCARDIAL INFARCTION, DISCHARGED ALIVE WITH CC |
| 291 | HEART FAILURE AND SHOCK WITH MCC |
| 255 | UPPER LIMB AND TOE AMPUTATION FOR CIRCULATORY SYSTEM DISORDERS WITH MCC |
| 283 | ACUTE MYOCARDIAL INFARCTION, EXPIRED WITH MCC |
| 252 | OTHER VASCULAR PROCEDURES WITH MCC |
| 258 | CARDIAC PACEMAKER DEVICE REPLACEMENT WITH MCC |
| 242 | PERMANENT CARDIAC PACEMAKER IMPLANT WITH MCC |
| 282 | ACUTE MYOCARDIAL INFARCTION, DISCHARGED ALIVE WITHOUT CC/MCC |
| 273 | PERCUTANEOUS INTRACARDIAC PROCEDURES WITH MCC |
| 250 | PERCUTANEOUS CARDIOVASCULAR PROCEDURES WITHOUT CORONARY ARTERY STENT WITH MCC |
| 192 | CHRONIC OBSTRUCTIVE PULMONARY DISEASE WITHOUT CC/MCC |
| 321 | PERCUTANEOUS CARDIOVASCULAR PROCEDURES WITH INTRALUMINAL DEVICE WITH MCC OR 4+ ARTERIES/INTRALUMINAL DEVICES |
| 276 | CARDIAC DEFIBRILLATOR IMPLANT WITH MCC OR CAROTID SINUS NEUROSTIMULATOR |
| 4 | TRACHEOSTOMY WITH MV >96 HOURS OR PDX EXCEPT FACE, MOUTH AND NECK WITHOUT MAJOR O.R. PROCEDURE |
| 194 | SIMPLE PNEUMONIA AND PLEURISY WITH CC |

**Supplementary Table 2:**
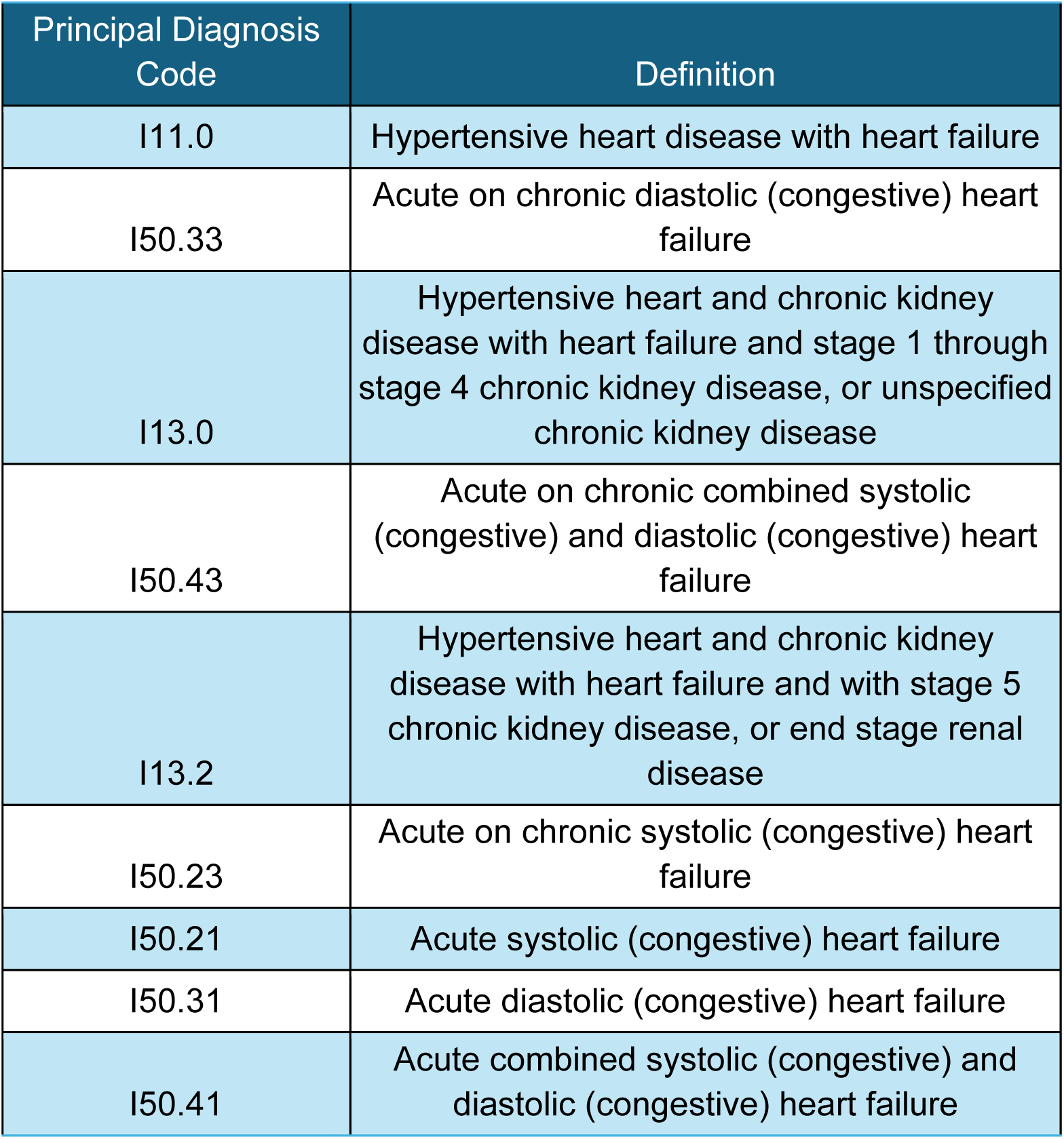
A List of Unique Principal Diagnosis Codes Used.

| Principal Diagnosis Code | Definition |
| --- | --- |
| I11.0 | Hypertensive heart disease with heart failure |
| I50.33 | Acute on chronic diastolic (congestive) heart failure |
| I13.0 | Hypertensive heart and chronic kidney disease with heart failure and stage 1 through stage 4 chronic kidney disease, or unspecified chronic kidney disease |
| I50.43 | Acute on chronic combined systolic (congestive) and diastolic (congestive) heart failure |
| I13.2 | Hypertensive heart and chronic kidney disease with heart failure and with stage 5 chronic kidney disease, or end stage renal disease |
| I50.23 | Acute on chronic systolic (congestive) heart failure |
| I50.21 | Acute systolic (congestive) heart failure |
| I50.31 | Acute diastolic (congestive) heart failure |
| I50.41 | Acute combined systolic (congestive) and diastolic (congestive) heart failure |

**Supplementary Table 3:** Chi Square Test of Independence Results for Marital Status and Sex (* indicates significance)

|  | Observed |  | Expected |  |
| --- | --- | --- | --- | --- |
| **Readmit** | 0 | 1 | 0 | 1 |
| **Married Female** | 136 | 9* | 125.839 | 19.161* |
| **Married Male** | 160 | 24 | 159.686 | 24.314 |
| **Single Female** | 334 | 56 | 338.464 | 51.536 |
| **Single Male** | 342 | 59 | 348.011 | 52.989 |

**Supplementary Table 4:** Dunn’s post hoc tests and Benjamini-Hochberg corrections for Chi-Square Test of Independence (* indicates significance)

| **<u>Benjamini-Hochberg Corrections</u>** |  |  |  |
| --- | --- | --- | --- |
| **Group** | **Outcome** | **P Value Uncorrected** | **P Value Corrected** |
| Married Female | 0 | 0.365 | 0.980 |
| Married Female | 1 | 0.020* | 0.162 |
| Married Male | 0 | 0.980 | 0.980 |
| Married Male | 1 | 0.949 | 0.980 |
| Single Female | 0 | 0.808 | 0.980 |
| Single Female | 1 | 0.534 | 0.980 |
| Single Male | 0 | 0.747 | 0.980 |
| Single Male | 1 | 0.409 | 0.980 |

**Table 5:**
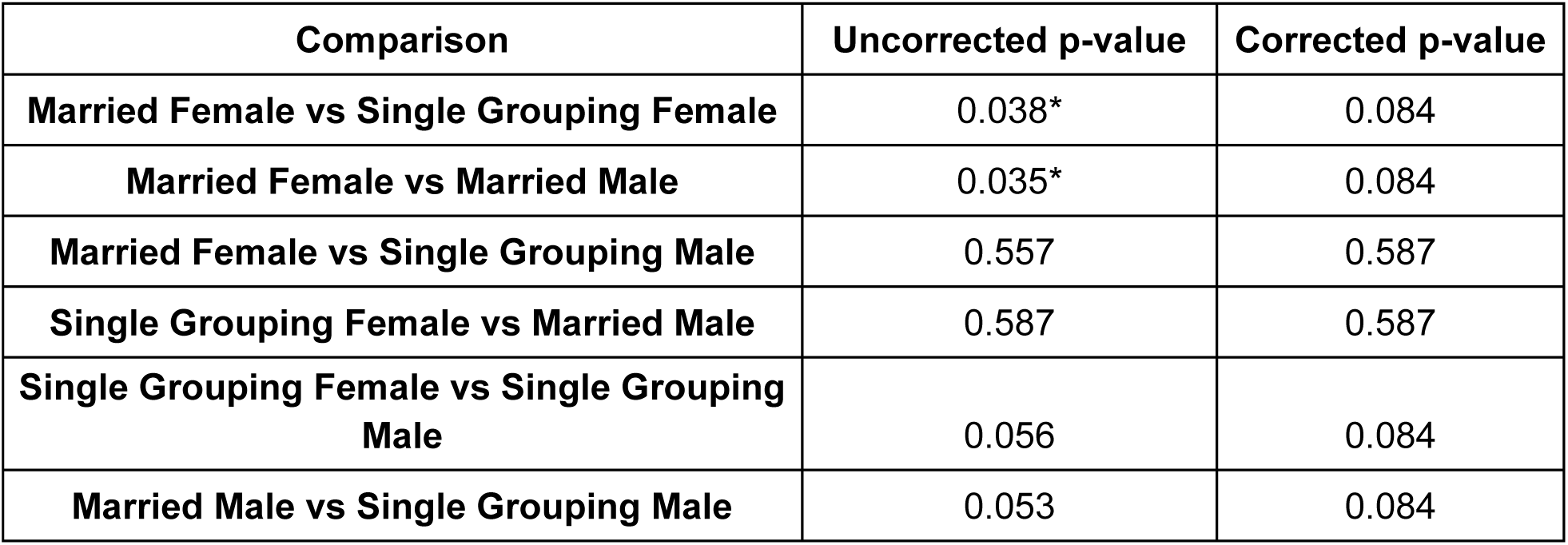
Post-hoc analyses for Kruskal Wallis Test for Total Length of Stay (* indicates significance)

| **Comparison** | **Uncorrected p-value** | **Corrected p-value** |
| --- | --- | --- |
| **Married Female vs Single Grouping Female** | 0.038* | 0.084 |
| **Married Female vs Married Male** | 0.035* | 0.084 |
| **Married Female vs Single Grouping Male** | 0.557 | 0.587 |
| **Single Grouping Female vs Married Male** | 0.587 | 0.587 |
| **Single Grouping Female vs Single Grouping Male** | 0.056 | 0.084 |
| **Married Male vs Single Grouping Male** | 0.053 | 0.084 |

**Table 6:** Odds Ratios and 95% Confidence Intervals (CI) for Readmission based on Marital Status and Sex Categories (* indicates significance)

|  | **odds ratio** | **coef** | **std err** | **z** | **P>\|z\|** | **[0.025** | **0.975]** |
| --- | --- | --- | --- | --- | --- | --- | --- |
| **const** | -- | -2.715 | 0.344 | -7.889 | 0.000* | -3.390 | -2.041 |
| **Married Male** | 2.248 | 0.818 | 0.408 | 2.006 | 0.045* | 0.019 | 1.618 |
| **Single Female** | 2.532 | 0.930 | 0.373 | 2.491 | 0.013* | 0.198 | 1.661 |
| **Single Male** | 2.606 | 0.958 | 0.372 | 2.576 | 0.010* | 0.229 | 1.687 |

